# Site-specific decrease in cortical reactivity during sensory trick in cervical dystonia patients

**DOI:** 10.1101/2021.02.01.21250820

**Authors:** Nivethida Thirugnanasambandam, Shivangi Singh, Hyun Joo Cho, Hitoshi Shitara, Pattamon Panyakaew, Sang Wook Lee, Mark Hallett

## Abstract

**Background:** Sensory tricks (SeT) are various maneuvers that can alleviate dystonic contractions and are a characteristic feature of cervical dystonia (CD). The neurophysiology underlying SeT, however, remains largely unknown. Reducing the abnormal cortical facilitation and modulating the abnormal cortical and subcortical oscillatory activity are mechanisms that have been proposed. The supplementary motor area (SMA) and primary sensorimotor cortices are thought to be relevant to this phenomenon.

**Objective:** In the current study, using concurrent EEG recording during transcranial magnetic stimulation (TMS) of the SMA and primary motor cortex (M1), we aimed at determining the changes in cortical reactivity and oscillatory changes induced by SeT.

**Methods:** We recruited 13 patients with CD who exhibited SeT and equal number of age- and gender-matched healthy controls. Single TMS pulses were delivered over the SMA and M1 either at rest or during SeT. 32-channel EEG was recorded, and TMS-evoked potentials (TEP) were obtained. Further, time-frequency analysis was performed on the induced data. Correlation analysis for significant neurophysiological parameters was done with clinical measures.

**Results:** We found that SeT induced a significant decrease in the amplitude of TEP elicited from M1 stimulation at ∼210-260ms in patients, which correlated with symptom duration. Post hoc analysis of EMG activity in the neck muscles revealed that this effect on TEP was present only in the subset of patients with effective SeT.

**Conclusion:** Our results suggest that SeT reduces cortical reactivity over M1 approximately 200ms after stimulation. This adds support to the idea that reduced cortical facilitation underlies the phenomenon.

## Introduction

Sensory trick (SeT) is a characteristic feature of cervical dystonia (CD), a disorder that manifests as painful, abnormal, involuntary contraction of the neck muscles resulting in twisting or turning of the head to one side [1]. SeT may be defined as an episodic and specific maneuver that temporarily relieves dystonia in a manner that is not physiologically perceived as necessary to counteract the involuntary movement [2]. The most common form of SeT involves the patient touching his or her chin with the hand contralateral to the direction of the head turn [3]. Most studies have reported about 70-80% of CD patients exhibiting SeT [2]. However, only a handful of studies have examined the neurophysiology underlying SeT. It is known from transcranial magnetic stimulation (TMS) studies that SMA facilitates motor cortex output [4] and hypometabolism in SMA induced by SeT as observed on positron emission tomography [5] would decrease the facilitatory input to the primary motor cortex. Accordingly, Amadio and colleagues [6] showed using TMS that CD patients exhibited higher intracortical facilitation compared to healthy controls and that SeT significantly reduced the abnormal facilitation, suggesting that reduced cortical facilitation could possibly underlie the phenomenon of SeT. Another mechanism that has been proposed based on local field potential recordings in CD patients is the abnormal increase in low frequency oscillatory activity (3-12Hz) in globus pallidus interna [7-11]. There is good evidence to suggest that abnormal slow wave activity in the GPi partly contributes to the pathophysiology of CD and that SeT modulates the abnormal slow wave oscillatory activity to reduce dystonia [12]. In the current study, we aimed to further examine the neurophysiology underlying SeT using concurrent TMS-EEG recording [13, 14]. By using this integrated approach, it is possible to record TMS-evoked potentials that reflect changes in cortical reactivity evoked by TMS pulses [15] and the induced oscillatory activity [16, 17]. Since SMA and primary motor cortex (M1) are thought to be relevant to the neurophysiological mechanism underlying SeT, we delivered TMS pulses to both these regions and aimed at determining the changes in cortical reactivity and oscillatory activity induced by SeT. Further, we also intended to determine if any of the relevant neurophysiological parameters correlated with clinical measures of CD.

## Methods

### Study participants

Thirteen CD patients (mean age: 59.4 ± 13.3 (SD) years; 7 females) and 13 age-and gender-matched healthy controls participated in the study (Table 1). All participants were right handed as assessed by the Edinburgh Handedness Inventory of Manual Preference (Oldfield, 1971). All participants were screened by a physician or nurse practitioner for eligibility to undergo TMS and magnetic resonance imaging (MRI). All patients had a diagnosis of CD confirmed by a movement disorders specialist and did not have any other neurological illness. They all demonstrated sensory tricks (i.e., gently touching their face improved their symptoms). Their last injection of Botulinum toxin was at least 11 weeks prior to the study. Patients were considered ineligible for participation if they had any abnormal neurological signs other than dystonia, any history of brain tumor, stroke or head trauma. Other exclusion criteria included the use of any medications, including benzodiazepines, anticholinergics and antidepressants, in the two weeks prior to the study. Healthy volunteers (HV) underwent general physical and neurological examination as part of the screening for participation. They were excluded if they had any current or history of neurological/psychiatric illness or had used any medication acting on the central nervous system in the last two weeks. All participants were asked to abstain from alcohol and caffeine for 48 hours prior to the study. The study was approved by the CNS Institutional Review Board of the National Institutes of Health and conformed to the ethical guidelines of the Declaration of Helsinki. Written informed consent was obtained from all participants prior to the study.

**Table 1.**
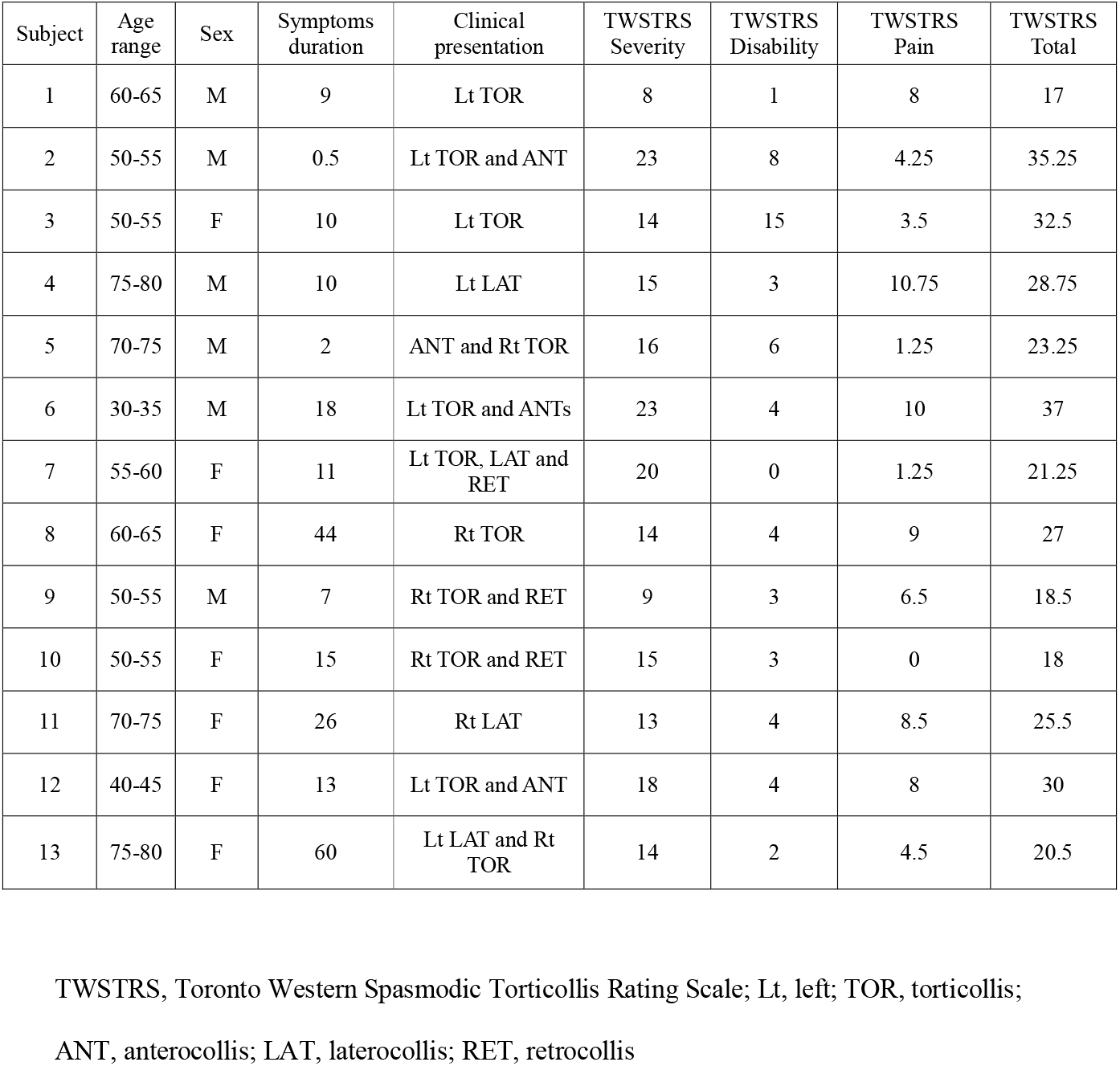
Clinical characteristics of patients with cervical dystonia.

| Subject | Age range | Sex | Symptoms duration | Clinical presentation | TWSTRS Severity | TWSTRS Disability | TWSTRS Pain | TWSTRS Total |
| --- | --- | --- | --- | --- | --- | --- | --- | --- |
| 1 | 60-65 | M | 9 | Lt TOR | 8 | 1 | 8 | 17 |
| 2 | 50-55 | M | 0.5 | Lt TOR and ANT | 23 | 8 | 4.25 | 35.25 |
| 3 | 50-55 | F | 10 | Lt TOR | 14 | 15 | 3.5 | 32.5 |
| 4 | 75-80 | M | 10 | Lt LAT | 15 | 3 | 10.75 | 28.75 |
| 5 | 70-75 | M | 2 | ANT and Rt TOR | 16 | 6 | 1.25 | 23.25 |
| 6 | 30-35 | M | 18 | Lt TOR and ANT | 23 | 4 | 10 | 37 |
| 7 | 55-60 | F | 11 | Lt TOR, LAT and RET | 20 | 0 | 1.25 | 21.25 |
| 8 | 60-65 | F | 44 | Rt TOR | 14 | 4 | 9 | 27 |
| 9 | 50-55 | M | 7 | Rt TOR and RET | 9 | 3 | 6.5 | 18.5 |
| 10 | 50-55 | F | 15 | Rt TOR and RET | 15 | 3 | 0 | 18 |
| 11 | 70-75 | F | 26 | Rt LAT | 13 | 4 | 8.5 | 25.5 |
| 12 | 40-45 | F | 13 | Lt TOR and ANT | 18 | 4 | 8 | 30 |
| 13 | 75-80 | F | 60 | Lt LAT and Rt TOR | 14 | 2 | 4.5 | 20.5 |

TWSTRS, Toronto Western Spasmodic Torticollis Rating Scale; Lt, left; TOR, torticollis;
ANT, anterocollis; LAT, laterocollis; RET, retrocollis

### Experimental procedure

Participants were seated comfortably on a chair with their forearms resting on a side table. An EEG cap (Braincap MR, Easycap, Munich, Germany) with 32 electrodes was positioned on their heads. Subjects also wore glasses with a subject tracker for neuronavigation whose position was secured throughout the experiment and was calibrated using the infrared camera of the optical neuronavigation system. Two Magstim200^2^ stimulators connected through a Bistim unit (Magstim Company, Whitland, Dyfed, UK) and to a figure-of-eight coil (70mm external loop diameter) were used to deliver TMS pulses. Stimulation was delivered either to M1 or to SMA contralateral to the resting hand at 110% of resting motor threshold intensity. Further details of the EMG, neuronavigation, TMS and EEG methods are provided in the supplementary information.

The experiment comprised of 8 blocks in total for each subject. We had two blocks (50 pulses/block) for each of the following conditions – M1_rest, M1_trick, SMA_rest, and SMA_trick, that is, 4 blocks were designated for each stimulation site. Altogether, 100 pulses were delivered over a stimulation site during either rest or SeT condition. Patients were asked to either stay in their resting dystonic position or perform the SeT that relieved the dystonia during the entirety of the respective block. Seven patients used their left hand for the SeT, while the others used their right hand. Controls were asked to mimic the rest and SeT head positions of their age- and gender-matched patient during the respective blocks. The investigator demonstrated the dystonic head position of the patient matched to every healthy volunteer. The order of the blocks was randomized across participants. One hundred single, monophasic TMS pulses were delivered over M1 or SMA contralateral to the resting hand (i.e., the hand that did not perform the SeT). Supplementary figure 1B shows an illustration of the experimental protocol. EEG data analysis was performed using the EEGLAB [18] and Fieldtrip [19] open source MATLAB toolboxes. A detailed description of the data analysis methods is provided in the supplementary information. We performed Spearman’s correlation analysis of the statistically significant EEG parameters with clinical measures - duration of symptoms, disease severity and disability as assessed by Toronto Western Spasmodic Torticollis Rating Scale (TWSTRS).

**Figure 1.**
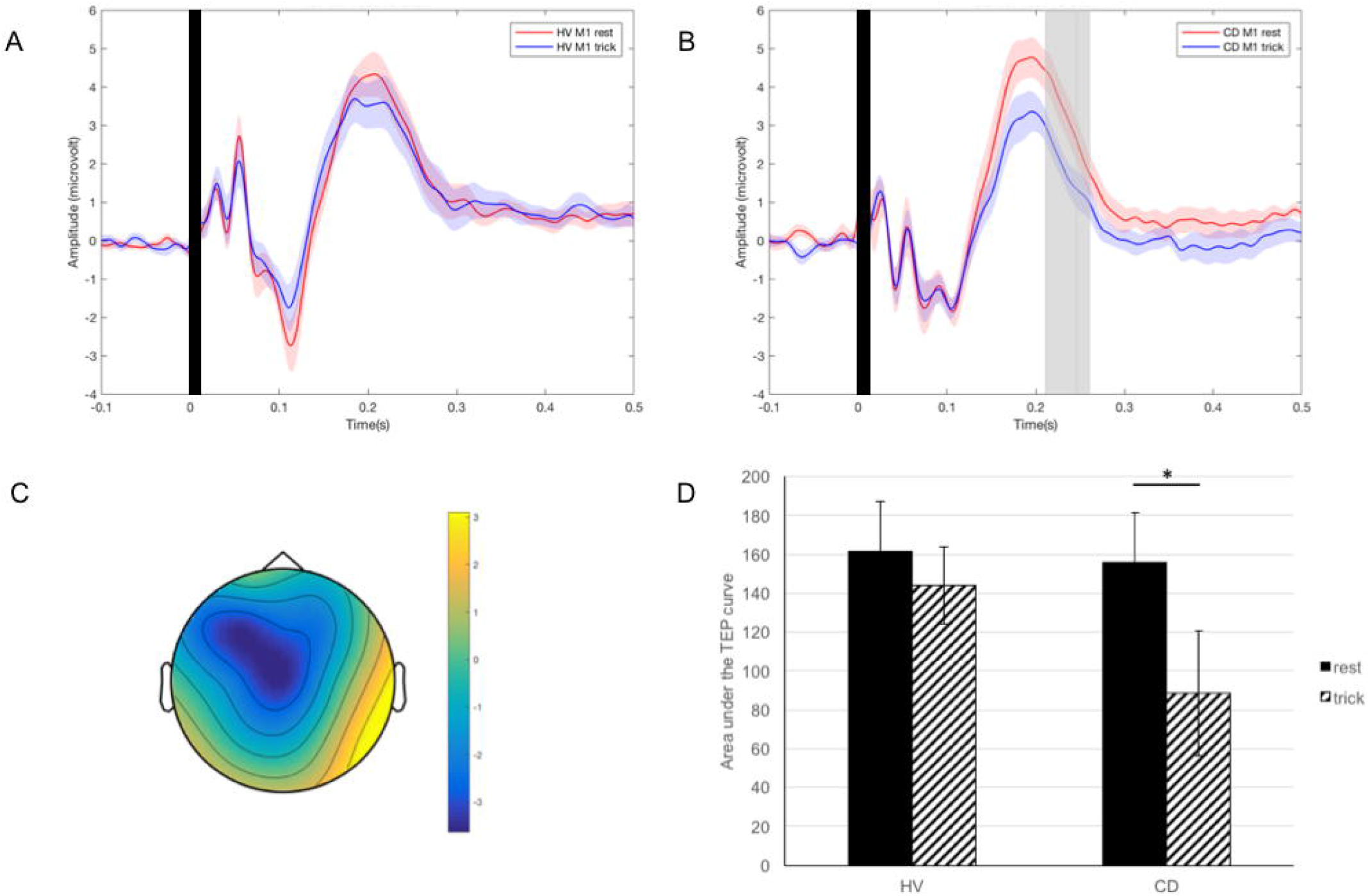
(A) TMS-evoked potential (TEP) in healthy controls at rest (red) and during trick (blue). (B) TEP in cervical dystonia patients at rest (red) and during trick (blue). TEPs were evoked by stimulation over M1. Time window showing statistically significant difference between rest and trick conditions is in grey. (C) Topographic map of the scalp localizing the difference in cortical activity between rest and trick conditions in patients. (D) Bar graph showing the mean area under the curve of the TEP within the significant time window in both conditions (black bars: rest; shaded bars: trick) and subject groups (HV: healthy controls; CD: cervical dystonia patients). Error bars are standard error of mean. Asterisk indicates p<0.05.

Post hoc analysis: EMG data from the right SCM (RSCM) and the left SCM (LSCM) muscles were band-pass filtered (0.5Hz – 100Hz) then a notch filter (60Hz) was applied to remove power-line noise. Temporal profile of the muscle activation was obtained by applying a low-pass filter (5Hz) to the rectified EMG signals. The relative change in the muscle activation during SeT was computed from the muscle activation during the baseline and during SeT; here, the log change in percent (L%; Δ_L_) was used to estimate the change in the SCM muscle activity (Δ_L_SCM), which resolves the problem of asymmetry and non-additivity of the relative measure [20].

## Results

All participants completed the experiment without any adverse event. The average number of trials included for the analysis were: 87 ± 5.97 for M1_rest, 87 ± 5.97 for M1_trick, 76 ± 12.42 for SMA_rest, and 76 ± 13.6 for SMA_trick.

### M1 stimulation

#### TEP analysis

The TEP in all groups and conditions showed a large positive peak at ∼200 ms latency, corresponding to the P180 component which is described in literature as the largest positive value between 130 and 230ms [21, 22]. Comparison of SeT vs rest in CD patients showed a smaller peak. A quantitative analysis of the difference, reflecting this reduction in positivity, revealed a significant negative cluster (p = 0.014) from 211-260ms over the fronto-central region including electrodes C3, FC1, Fz, Cz and FC2 (Figure 1). Post hoc analysis (described below) revealed that the reduction in P180 amplitude induced by SeT was seen only in the subgroup of patients that exhibited effective SeT. A similar comparison in HVs did not reveal any significant difference between the 2 conditions. No significant clusters were obtained by comparing the baseline at rest or the change induced by SeT in CD patients with that in HVs.

#### Time-frequency analysis

No significant differences in oscillatory power were observed between conditions in either of the subject groups (Supplementary figure 2). A between-groups comparison at rest also did not reveal any significant difference.

**Figure 2.**
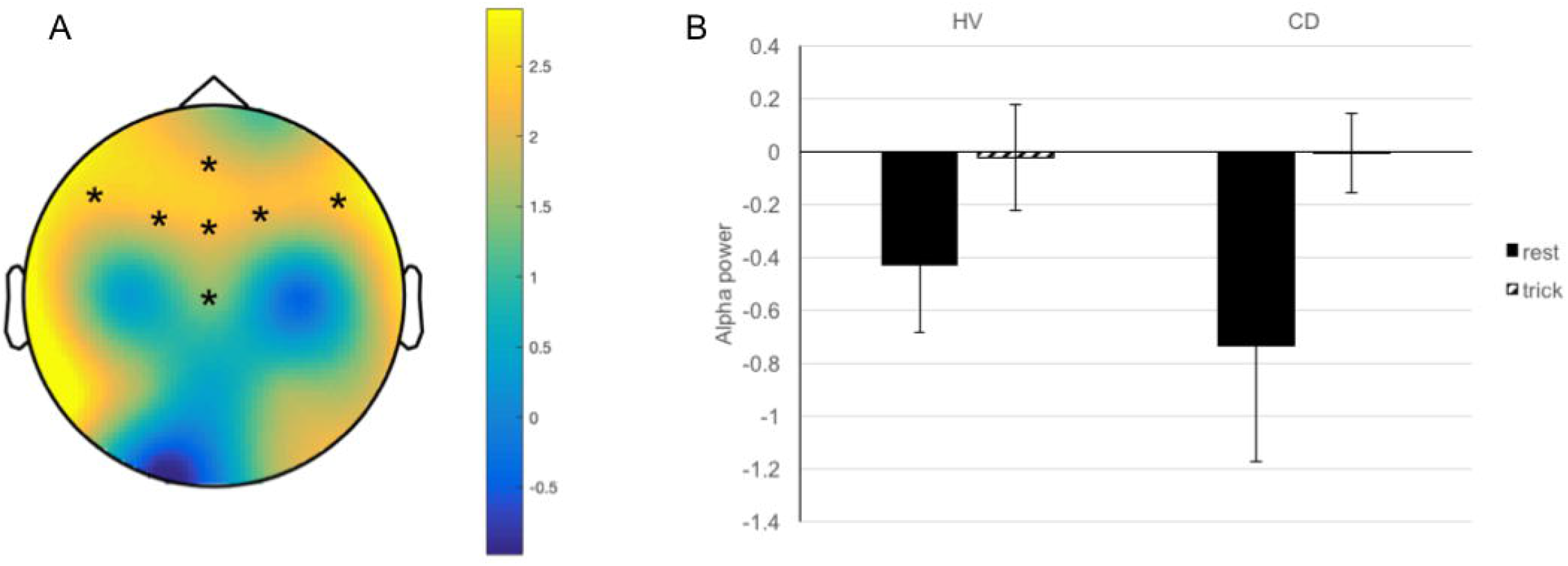
(A) Topographic map of the scalp showing the significant positive cluster of electrodes (Fz, FCz, Cz, FC1, FC2, FC5 and FC6), that is, electrodes where there was significant increase in alpha power during trick in patients with cervical dystonia for SMA stimulation. (B) Bar graph showing the mean alpha power during the significant time window in both conditions (black bars: rest; shaded bars: trick) and subject groups (HV: healthy controls; CD: cervical dystonia patients) for SMA stimulation.

### SMA stimulation

#### TEP analysis

No significant differences in TEPs were observed between conditions or subject groups (Supplementary figure 3).

**Figure 3.**
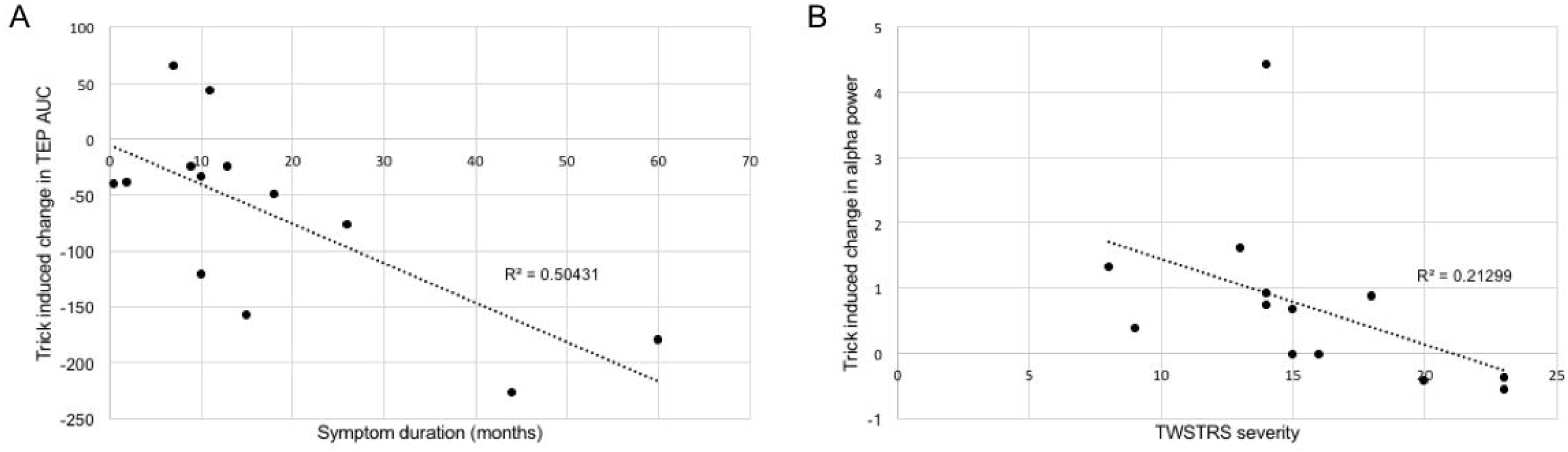
Correlation analysis (A) between symptom duration and trick-induced change in area under the curve of TMS-evoked potential within the significant time window (B) between TWSTRS severity score and trick-induced change in alpha power within the significant time window

#### Time-frequency analysis

Comparison of the SeT and rest conditions in CD patients revealed a significant positive cluster (p=0.02) in the alpha band at 500ms over the fronto-central region bilaterally including electrodes FCz, FC1, FC5, Cz, Fz, FC2 and FC6 (Figure 2). Post hoc analysis (described below) showed that the change in alpha power was similar in both subgroups of patients, irrespective of whether the SeT was effective or forced. A similar comparison of SeT and rest conditions in HVs did not yield any significant cluster. Also, between-groups comparison at rest did not reveal any significant difference.

#### Correlation analysis

We observed a significant positive correlation (p = 0.026) between the reduction in TEP amplitude induced by SeT and symptom duration. Also, trick-induced alpha synchronization was negatively correlated with disease severity (p = 0.003).

#### Post hoc analysis

EMG data from the SCM muscle were analyzed in all patients and we found that only in 6 out of 13 patients, the increase in the muscle activation was significantly smaller during SeT (Δ_L_SCM: mean±SD = 2.3±31.5% for dystonic side; 22.9±70.6% for contralateral side) than when they moved the neck without performing the trick (Δ_L_SCM: mean±S D = 164.5±187.1% for dystonic side; 68.3±130.1% for contralateral side). This implies that these patients exhibited an effective SeT. The remaining 7 patients showed similar degree of increase in the SCM muscle activity during the SeT (Δ_L_SCM: mean±SD = 57.4±117.3% for dystonic side; 18.0±25.5% for contralateral side) and during active neck movement (Δ_L_SCM: mean±SD = 61.8±135.4% for dystonic side; 22.6±29.9% for contralateral side), thus revealing that they exhibited a ‘forcible’ trick [2]. Supplementary figure 4 shows the EMG data from a representative subject from each of the 2 subgroups.

## Discussion

The results of our study clearly show that TMS-evoked responses are site-specific. We did not observe any significant group difference in either evoked or induced response to SMA or M1 stimulation at rest. Phase-locked responses to M1 stimulation resulted in significantly smaller P180 in CD patients during SeT. Notably, the reduction in P180 amplitude over M1 correlated positively with symptom duration. Although SMA stimulation did not modulate the TEP, it revealed a significant difference in TMS-induced oscillatory response during SeT in CD patients. We found a significant increase in alpha power at 500ms spread over the fronto-central electrodes bilaterally. Post hoc analysis revealed that the P180 amplitude reduction was associated with an effective SeT while the change in alpha activity was non-specific.

### Impact of SeT on TMS-evoked response

In the current study, we have shown that SeT in CD patients significantly decreased the amplitude of the long-latency positive component of the TEP – the P180. The neural basis of the different TEP components is still remains unclear [13]. Many studies suggest that the N100-P180 complex is contaminated by the auditory and somatosensory evoked responses, particularly bone conducted sound [23, 24]. Nevertheless, there is evidence to suggest that P180 also contains significant cortical activity evoked by the TMS pulse [15, 25, 26] and hence it cannot be ignored as artefactual. It is thought that P180 may represent a late excitatory component [27, 28]. Our results are most consistent with a change in cortical activity induced by SeT modulating the P180 amplitude. It is unlikely that this effect is solely due to bone conduction because we compared the TEPs elicited at the same stimulation intensity in all conditions, and the P180 amplitude was affected only following M1 stimulation in the patients. The site specificity of the effect does not support the purely auditory origin of P180. Moreover, post hoc analysis revealed that the P180 reduction was seen exclusively in those patients who exhibited an effective SeT and not in those who had a forcible trick. This reinforces that the reduction in P180 amplitude could be primarily due to change in cortical activity induced by SeT. TMS studies have demonstrated that CD patients have high intracortical facilitation, which is reduced by SeT [6]. Considering that P180 represents an excitatory component of TEP, our results also suggest that reduction in cortical facilitation could underlie an effective SeT.

### Impact of SeT on TMS-induced oscillations

Contrary to our expectations, we did not find any significant effect of SeT on TMS-induced oscillations. Although we observed an increase in alpha power that was significant in CD patients, post hoc analysis revealed that this might not be relevant to the neurophysiology of SeT since this effect was observed irrespective of the effectiveness of SeT. Further, HVs who mimicked the head posture of the patients also showed similar trend. Figure 3 shows that both HVs and patients had low alpha power in the dystonic posture (alpha desynchronization). Changing the head position to neutral relaxed the neck muscles and thereby could have abolished this desynchronization. This seems likely because the SeT-induced change in alpha power correlated negatively with disease severity. That is, those who had severe disease, had stronger contraction of the neck muscles even at neutral head position achieved by SeT and hence less change in alpha power. Based on past studies, we expected that SeT would cause desynchronization of low frequency bands especially in the theta range [7]. There has been recent evidence showing that pallidal theta may be significant in the pathophysiology of CD [9]. CD patients showed high pallidal theta activity and SeT caused theta desynchronization in them. Further, Tang et al. [3] also demonstrated that pallidal and thereby cortical theta desynchronization could underlie an effective SeT. However, we did not observe any changes in the cortical theta activity. We think that it could be because we recorded EEG during tonic phase of SeT. The trick was maintained for about 5 minutes until 50 TMS pulses were delivered at 5-7s intervals within a block. Tang and colleagues recorded theta desynchronization about 4-6s following the onset of SeT. Due to our experiment design, we could have missed out on this observation.

## Conclusion

In the current study, we have shown that integrating TMS and EEG can be a very useful multimodal approach to explore physiological phenomena that are particularly altered in disease states. One major technical limitation of the study is that we did not use white noise or any foam padding to reduce the impact of auditory or somatosensory stimulation from the TMS on the TMS-evoked potentials. Although it would have been ideal to have used these measures, we think that these confounds are unlikely to have affected our results. Our results clearly demonstrate that site-specific cortical effects can also be observed with long-latency components of TEPs and may be of relevance to disease pathophysiology. Thus, concurrent TMS-EEG could be a promising tool in elucidating the pathophysiology of neuropsychiatric disorders.

## Data Availability

Data will be made available on the NIH repository after the acceptance of manuscript for publication.

## Acknowledgements

This study was funded by NINDS intramural research program.

## Author roles

1. Research project:
  a. Conception: NT, HJC, MH
  b. Organization: NT, HJC
  c. Execution: HJC, HS, PP, SS
2. Statistical analysis:
  a. Design: NT; SWL
  b. Execution: SS; NT; SWL
  c. Review and Critique: NT, MH
3. Manuscript:
  a. Writing of the first draft: NT, SS
  b. Review and Critique: HJC, HS, PP, SWL, MH

