## Supplementary_information for "Site-specific decrease in cortical reactivity during sensory trick in cervical dystonia patients"

### METHODOLOGY

#### Neuronavigation

Individual anatomical MRI images (from 3T MR scanner) was used to localize SMA and monitor the coil position throughout the course of the experiment. MRI-guided optical neuronavigation system (Brainsight TMS, Rogue Resolutions, Cardiff, UK) combined with a tracking system ("Polaris Vicra", Northern Digital Inc., Waterloo, Canada) was used to identify the site of stimulation. SMA was identified as the area just posterior to the vertical line passing through the anterior commissure and perpendicular to the AC-PC plane [1].These sites were marked as targets on the Brainsight software and were monitored throughout the experiment such that the spatial error was always limited to less than 2mm.

#### EMG recording

Bipolar Ag/AgCl adhesive electrodes were placed on the skin over the first dorsal interossous (FDI) of the resting hand and bilateral sternocleidomastoid (SCM) muscles. The EMG signal was recorded, amplified and filtered (20 Hz-1 kHz bandwidth) using a Nihon Kohden Neuropack MEB 4300 system (Nihon Kohden Corporation, Tokyo, Japan) and was digitized at a sampling frequency of 5 kHz with a CED micro1401 A/D laboratory interface (Cambridge Electronic Design, Cambridge, UK). Data were stored in a laboratory computer for offline analysis using Signal v6.1 (Cambridge Electronic Design, Cambridge, UK).

#### Transcranial magnetic stimulation

Two Magstim200$^2$ stimulators connected through a Bistim unit (Magstim Company, Whitland, Dyfed, UK) and to a figure-of-eight coil (70mm external loop diameter) were used to deliver TMS pulses. Stimulation was delivered either to M1 or to SMA contralateral to the resting hand. For M1 stimulation, the coil was positioned at an approximate angle of 45° from the midline so as to induce a postero-anterior current in the brain, perpendicular to the central sulcus. The motor 'hotspots' for FDI and SCM were identified as the points on the scalp which evoked the largest motor evoked potentials (MEP) from the respective muscle consistently. The resting motor threshold (RMT) was recorded from the FDI as the minimum stimulation intensity required to elicit an MEP of 50 microvolt amplitude in at least 50% of trials using adaptive threshold hunting procedure [2]. For M1 stimulation, the coil was positioned over the motor hotspot for SCM and monitored constantly using neuronavigation. For SMA stimulation, the stimulation site was identified using the anatomical MRI of the individual subject and the coil was positioned perpendicular to the sagittal sulcus with the handle pointing outwards so as to induce a medially directed current in stimulated cortex. Stimulation intensity equal to 110%RMT was used for both M1 and SMA stimulations.

### EEG recording

EEG was recorded from 32 channels using a TMS compatible EEG system (Brainamp MR, Brain Products GmbH, Munich, Germany), which prevents saturation of the EEG amplifier by the TMS pulse and allows for continuous data recording. The scalp beneath the electrodes was prepared using an abrasive and conductive gel to achieve an impedance of less than 5 kΩ throughout the recording. The EEG signal was sampled at 5 kHz and a resolution of 0.1 μV/div. Supplementary figure 1A shows the electrode cap layout. We used AFz and Pz as the ground and reference electrodes, respectively. During the entire study, patients were asked to keep their eyes open and remain alert. Earplugs were used to reduce contamination of the EEG signal by acoustic stimuli. Unfortunately, we did not use any noise masking and consider this to be one of the limitations of our study, which we have discussed in detail later.

### EEG data analysis

**Preprocessing:** Data analysis was performed using the EEGLAB [3] and Fieldtrip [4] open source MATLAB toolboxes. EEG data were segmented into trials from -0.5 to 1s centered around the TMS pulse. Trials were inspected manually and those with excessive artifacts (excessive muscle contraction, bad electrode contact, gross head/neck movement) were removed. The TMS pulse artifact was removed by excluding data between -1 to 6ms around the TMS pulse. Independent Component Analysis was performed and an average of 5 components corresponding to eye blinks, eye movements and decay artifact were removed. A liberal window of 0-20ms that included the muscle artifact was also rejected. Cubic interpolation of the rejected data points was performed. The data were re-referenced to the common average reference, bandpass filtered to 2-50Hz and downsampled to 1KHz. EEG trials were then visually scrutinized, and those that still contained artifacts that may have been missed through previous methods were eliminated. Finally, baseline correction was done by subtracting the mean baseline amplitude during a time window between 100ms and 10ms prior to the TMS pulse. Artifact-free trials were then averaged (approximately 80 trials per block) to obtain TMS-evoked potentials (TEP).

**TEP analysis:** We used a data-driven method to identify electrode clusters that were significantly different between groups/conditions. We made 3 comparisons for each stimulation site (M1 and SMA) – (i) SeT vs rest in CD patients (ii) SeT vs rest in healthy controls and (iii) rest or baseline activity in CD patients vs controls. Non-parametric cluster-based permutation analysis [5] was performed for each of the above comparisons to identify the time windows and the electrode clusters that showed statistically significant difference between the groups/conditions compared. Electrodes that constituted the significant clusters were noted and TEPs recorded from them were averaged and plotted for illustration. Area under the curve was calculated from the averaged TEP of the cluster for the time window that showed significant difference and used for correlation analysis with clinical measures.

**Time-frequency analysis:** Time frequency analysis was performed on the induced data to explore the impact of the TMS pulse on the power spectrum of different frequency bands (theta: 4-7Hz, alpha: 8-13Hz, beta: 14-30Hz and gamma: 31-50Hz). To obtain the induced data, we subtracted the TEP from individual trials of each subject and condition [6]. Time frequency representations were calculated using short-time Fourier transforms with Hanning window tapering. Sliding time windows of 200ms length were used with 50% overlap between consecutive segments, yielding a frequency resolution of approximately 1Hz. Spectral power was calculated for all frequency bins from 2Hz to 50Hz (comprising theta, alpha, beta and gamma bands) for each subject and condition by averaging the respective values across all trials. We performed non-parametric cluster-based permutation analysis [5] for each frequency band to identify the time points and electrode clusters that showed significant difference between groups/conditions. Three comparisons were made for each frequency band as described earlier for the TEPs. Finally, the spectral power over electrodes that constituted the significant clusters were averaged across all frequencies within a specific band and time window for every subject and condition. This parameter was used for correlation analysis with clinical measures.

**Supplementary Figure 1. (A)** Layout of electrode cap. Pz = Reference electrode; AFz = Ground electrode. **B.** Illustration of experimental protocol. Upper panel: Stimulation of motor hotspot for sternocleidomastoid along the postero-anterior direction (i) in rest condition when head is in dystonic posture and (ii) during sensory trick when dystonia is relieved. Lower panel: Stimulation of supplementary motor area along the latero-medial direction (iii) in rest condition when the head is in dystonic posture and (iv) during sensory trick when dystonia is relieved. A hundred TMS pulses divided into 2 equal blocks were delivered for each of the 4 conditions in a random order for each subject.

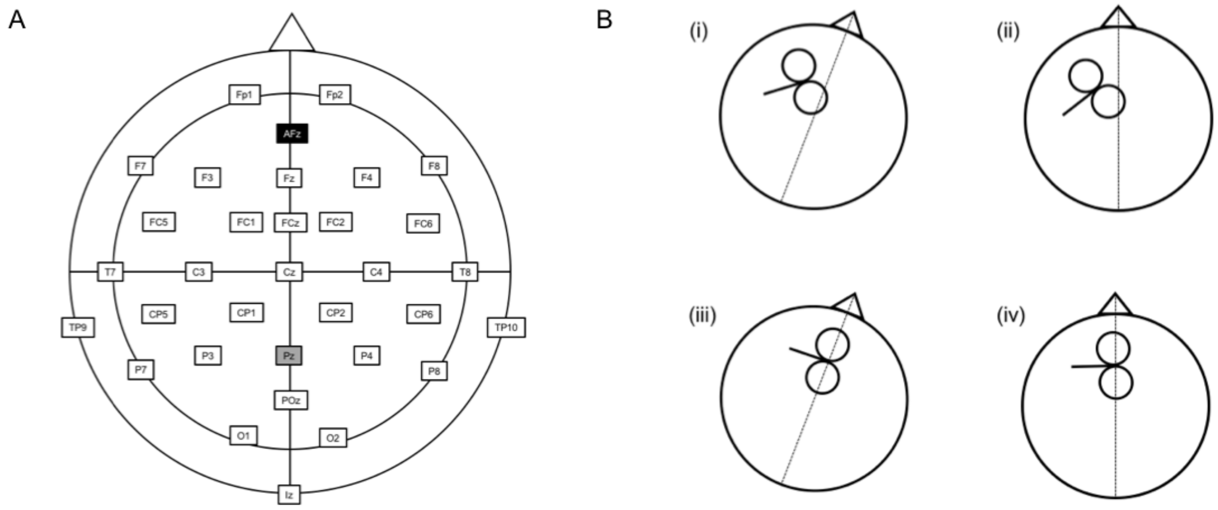

**Supplementary Figure 2:** Bar graph showing the mean alpha power during in both conditions (black bars: rest; shaded bars: trick) and subject groups (HV: healthy controls; CD: cervical dystonia patients) for M1 stimulation.

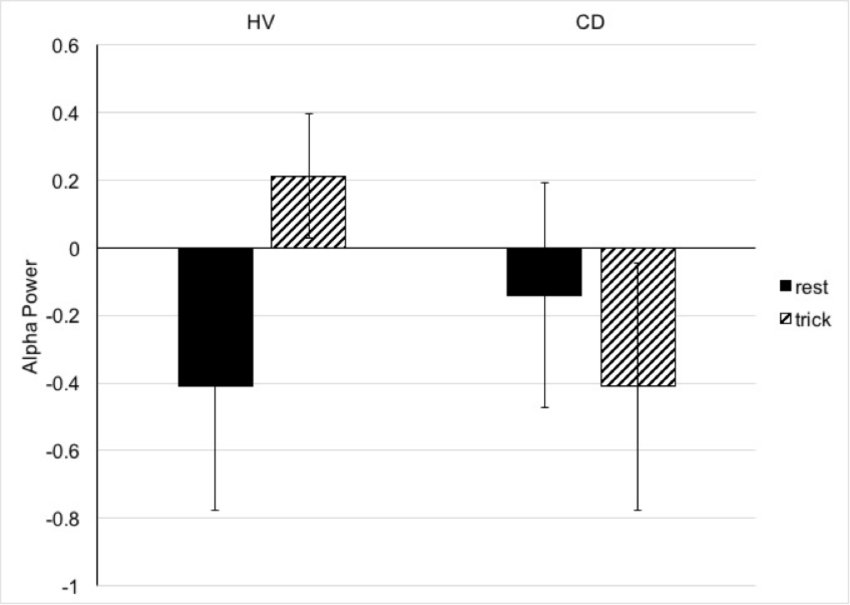

**Supplementary Figure 3:** Left: TMS-evoked potential (TEP) in healthy controls at rest (red) and during trick (blue). Right: TEP in cervical dystonia patients at rest (red) and during trick (blue). TEPs were evoked by stimulation over SMA.

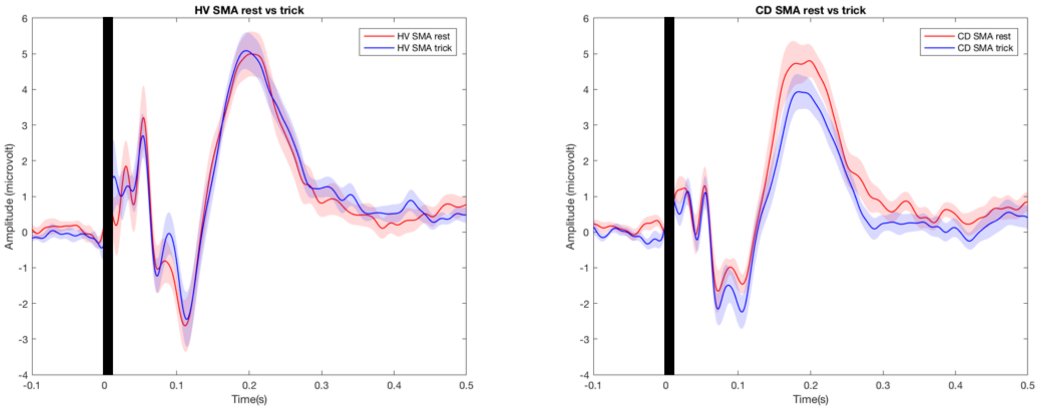

**Supplementary Figure 4:** EMG activity recorded in the left (LSCM) and right (RSCM) sternocleidomastoid muscles during sensory trick and forced neck movement for 2 subjects representing each of the subgroups.

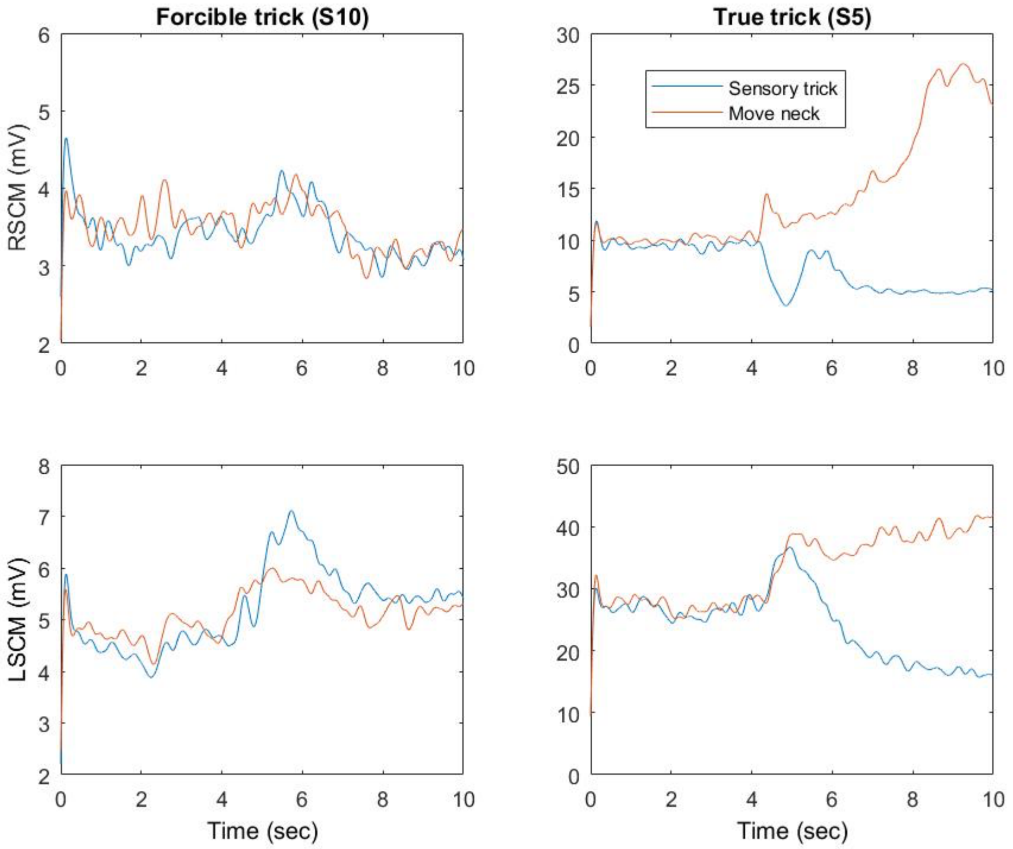
